# Nature and Trends of Pharmaceutical Payments to the Board Certified Respiratory Specialists in Japan between 2016 and 2019

**DOI:** 10.1101/2022.01.16.22269188

**Authors:** Anju Murayama, Momoko Hoshi, Hiroaki Saito, Sae Kamamoto, Manato Tanaka, Moe Kawashima, Hanano Mamada, Eiji Kusumi, Binaya Sapkota, Sunil Shrestha, Rajeev Shrestha, Divya Bhandari, Toyoaki Sawano, Erika Yamashita, Tetsuya Tanimoto, Akihiko Ozaki

## Abstract

**Background:** There are financial relationships between healthcare professionals and pharmaceutical companies, and these relationships have historically caused conflicts of interest and unduly influenced patient care. However, little was known about such relationship and its effect in clinical practice among specialists in respiratory medicine.

**Methods:** Based on the retrospective analysis of payment data made available by all 92 pharmaceutical companies in Japan, this study evaluated the magnitude and trend of financial relationships between all board-certified Japanese respiratory specialists and pharmaceutical companies between 2016 and 2019. Magnitude and prevalence of payments for specialists were analyzed descriptively. The payment trends were assessed using the generalized estimating equations for the payment per specialist and the prevalence of specialists with payments.

**Results:** Among all 7,114 respiratory specialists certified as of August, 2021, 4,413 (62.0%) received a total of USD($) 53,547,391 and 74,195 cases from 72 (78.3%) pharmaceutical companies between 2016 and 2019. The mean±SD and median (interquartile range) four-year combined payment values per specialist were $12,134±$34,045 and $2,210 ($715□$8,178) respectively. At maximum, one specialist received $495,332 personal payments over the four years. Both payments per specialist and prevalence of specialists with payments significantly increased during the four-year period, with 7.8% (95% CI: 5.5□9.8; p<0.001) in payments and 1.5% (95% CI: 0.61% □2.4%; p = 0.001) in prevalence of specialists with payments, respectively.

**Conclusion:** Majority of respiratory specialists had increasingly received substantial personal payments from pharmaceutical companies for the reimbursement of lecturing, consulting, and writing between 2016 and 2019. These increasing financial relationships with pharmaceutical companies might cause conflicts of interest among respiratory physicians.

## Introduction

Financial relationships between healthcare professionals and pharmaceutical companies might cause financial conflicts of interest (COIs) and jeopardize patient-centred care. Indeed, financial relationships between healthcare sectors and pharmaceutical companies might unintentionally influence physician prescribing patterns,^1-4^ recommendations of clinical guidelines,^5-9^ and conducting clinical research.^10-14^ Therefore, proper management of COIs is currently one of the most fundamental challenges for all healthcare professionals.^15^ To increase transparency in financial COIs in healthcare, Japan Pharmaceutical Manufacturers Association (JPMA), the largest pharmaceutical trade organization in Japan, mandate all member companies to publish all payments made to the physicians, including those for lecturing, writing, and consulting, itemizing the value of payments along with individuals’ names and affiliations on each company webpage from 2013 onwards, similar to the US Sunshine Act.^16^

Among various specialties, respiratory medicine is one of the increasing markets for pharmaceutical companies alongside improvement in diagnostic technology and increase in patients with respiratory diseases such as asthma. The market size for respiratory medications is forecasted to increase from 38,985 million USD in 2019 to 57,097 million USD in 2026 worldwide, and 2,519 million USD in 2019 to 3,160 million USD in 2026 in Japan.^17^ Further, lung cancer is one of the most common cancer types in Japan, with 125,636 cases prevalent per year (12.8% of all cancer patients) in 2018, ranking third out of 25 cancer types, and 75,394 annual deaths, ranking first out of 25 cancer types in 2019.^18^ The launch of mainly novel drugs for lung cancer contributed to the treatment of lung cancer and fierce market competition in Japan.^17^ Consequently, the payments from pharmaceutical companies are expected to be increasingly concentrated on respiratory specialists in Japan, as did in other specialities such as hematology. However, no studies have assessed the magnitude or trends in payments from pharmaceutical companies to respiratory specialists in Japan.

This study aimed to elucidate the prevalence of the board certified respiratory specialists receiving payment from pharmaceutical companies, the magnitude and trend of payments throughout recent years i.e., 2016-2019.

## Methods

### Participants

This retrospective study considered all of the Japanese Respiratory Society (JRS)-certified respiratory specialists certified as of August 12, 2021. The JRS (hereafter named as the Society) was established in 1961, and is the primary and the largest medical professional society in respiratory medicine in Japan, with 12,976 Society members as of January 2021. The Society contributed to training, certifying the respiratory physicians or specialists in Japan, with a name list of the specialists publicly disclosed on the Society webpage, development of clinical practice guidelines for respiratory diseases, promoting research and continuing medical educations (CME) in respiratory medicine, and publishing several academic journals such as the *Respiratory Investigation* and *Respirology* in English, and the *Annals of the Japanese Respiratory Society* in Japanese.

### Certification of respiratory specialists in Japan

To become a respiratory specialist in Japan, physicians had to complete all of the following seven criteria: (1) should be board-certified in internal medicine by the Japanese Society of Internal Medicine (JSIM) or surgery by the Japan Surgical Society (JSS); (2) should complete specialized training in respiratory medicine for at least 3 years at an institution accredited by the JRS after certification of an internal medicine specialist by the JSIM or a surgery specialist by the JSS; (3) must be a registered member of the JRS for at least three years; (4) must have three or more academic articles concerning respiratory medicine; (5) must attend clinical training course held by the JRS; (6) must be a non-smoker; and (7) must pass the written examination targeted for the respiratory specialist held by the JRS. Respiratory specialists need to renew their specialist certification every five years.

### Data collection

First, the name, affiliations, certification in teaching respiratory medicine, and address of all respiratory specialists were collected from the webpage of the JRS (https://www.jrs.or.jp/modules/senmoni/) on November 6, 2021. The payment data of JRS-certified respiratory specialists were extracted from all 92 pharmaceutical companies affiliated to the JPMA and other associated companies from 2016 to 2019 (19-21). The extracted data included recipients’ names and affiliations, monetary amount (in USD), number of payment cases, payment category, and pharmaceutical companies’ names. To remove payment data of different persons with similar names, we compared affiliations and recipients’ special-ties between the data from the Society and the pharmaceutical companies. For payments to specialists whose affiliation and specialty reported by pharmaceutical companies differed from those reported by the JRS, we manually searched the name of respiratory specialists and collected additional data from official institutional web pages to verify whether they were the same persons, similar to previous studies ^19,20^

According to the JPMA transparency guideline, only the payment concerning lecturing, writing, and consulting was disclosed along with individuals’ names and affiliations and analysed based on individuals’ specialists. Other types of personal payments such as meal, education, travel, and accommodations were not disclosed individually but only aggregated monetary value of these payments was disclosed by each company in Japan.^16^ Personal payments such as lecturing, writing, and consulting are directly and widely paid to physicians by pharmaceutical companies.^3,21,22^ Considering such nature of payments, the personal payments concerning lecturing, writing, and consulting were included in this study.

### Statistical Analysis

Descriptive analyses of payment values and the number of cases were performed based on the specialists and the pharmaceutical companies. As a few companies such as Shire Japan and Baxalta did not disclose the number of payment cases, the payments from these companies were excluded from the analysis for payment cases. The Gini index and the shares of the payment values held by the top 1%, 5%, 10%, and 25% of specialists were used to examine the payment concentration. The Gini index measures income inequality among a given population, ranging from 0 to1. The greater Gini index indicates a greater disparity in the distribution of payments on the specialist basis.^23-26^ Subgroup analyses were conducted to elucidate the geographical differences on prefecture and region level in the payments.

As the distribution of payments were highly right-skewed, the population-averaged generalized estimating equation (GEE) negative binomial regression model for the trend of payment value per specialist, and the linear GEE model log linked with binomial distribution for the trend of numbers of specialists with payments were used to evaluate payment trends through the four years. In addition, the relative percentage of the average annual increase in payments per specialist and in the prevalence of specialists with payments were used to report the results. The year of payments, and the proportion of physicians receiving payments and payment values were set as independent variable and dependent variable, respectively.^27-29^ Among data from 92 companies, 18 lacked payment data over the four years, because some companies were disaffiliated from the JPMA and others newly joined the JPMA. Therefore, we calculated only the payment trends of companies who had complete data of four-year. A two-sided p < 0.05 was considered statistically significant.

To adjust inflation and compare the payments across the study period, the Japanese yen (¥) was converted into the corresponding USD ($) using the 2019 average monthly exchange rate of ¥109.0 per $1. All analyses were conducted using Microsoft Excel, version 16.0 (Microsoft Corp), and Stata version 15 (StataCorp).

### Ethical approval

The Ethics Committee of the Medical Governance Research Institute approved this study (approval number: MG2018-04-20200605; approval date: June 5, 2020). As this study was a retrospective analysis of publicly available information from pharmaceutical companies and the Society webpage, informed consent was waived by the Ethics Committee.

## Results

A total of 7,114 respiratory specialists were certified by the JRS as of August 12, 2021, including 3,002 (42.2%) certified teaching respiratory specialists.

### Overview of payments

Our payment database recorded 1,474,653 payment cases worth $996,291,009 (¥108,595,720,027) from 92 pharmaceutical companies between 2016 and 2019. Of the 7,114 specialists, 4,413 (62.0%) received a total of $53,547,391 (¥5,836,665,622) and 74,195 payment cases from 72 (78.3%: 72 out of 92) pharmaceutical companies over the same period **(Table 1)**. The payment cases and values to the respiratory specialists occupied 5.0% and 5.4% of total cases and values, respectively. The mean±SD and median (interquartile range or IQR) four-year combined payment values per specialist were $12,134±$34,045 and $2,210 ($715□$8,178), respectively. At maximum, one specialist received $495,332 personal payments over the four years. The mean±SD and median (IQR) number of cases over the four years were 16.8±34.9 and 5.0 (2.0[16.0) cases per specialist. The respiratory specialists received the payments from 4.8±4.3; median: 3.0; and IQR: 1.0[7.0) pharmaceutical companies on average. Maximum 461 payment cases and 30 pharmaceutical companies per specialist were considered between 2016 and 2019.

**Table 1.** Summary of personal payments from pharmaceutical companies to respiratory specialists certified by the Japanese Respiratory Society between 2016 and 2019.

| **Variables** |  |
| --- | --- |
| **Total payments, \$** | |
| Payment values, \$ | 53,547,391 |
| Cases, n | 74,195 |
| Companies, n | 72 |
| **Average per specialist±SD** |  |
| Payment values, \$ | 12,134±34,045 |
| Cases, n | 16.8±34.9 |
| Companies, n | 4.8±4.3 |
| **Median (IQR)** |  |
| Payment values, \$ | 2,210 (715□8,178) |
| Cases, n | 5.0 (2.0□16.0) |
| Companies, n | 3.0 (1.0□7.0) |
| **Range** |  |
| Payment values, \$ | 31□495,332 |
| Cases, n | 1.0□461 |
| Companies, n | 1.0□30 |
| **Physicians with specific payments, n (%)** |  |
| Any payments | 4,413 (62.0) |
| Payments >\$500 | 3,747 (52.7) |
| Payments >\$1,000 | 3,000 (42.2) |
| Payments >\$5,000 | 1478 (20.8) |
| Payments >\$10,000 | 940 (13.2) |
| Payments >\$50,000 | 239 (3.4) |
| Payments >\$100,000 | 111 (1.6) |
| **Gini index** | **0.871** |
| **Category of payments, \$ (%)** | |
| Lecturing | 45,847,076 (85.6) |
| Consulting | 5,770,483 (10.8) |
| Writing | 1,701,141 (3.2) |
| Other | 228,691 (0.43) |

IQR: interquartile range; SD: standard deviation

**Table 2.** Trend of personal payments from pharmaceutical companies to respiratory specialists certified by the Japanese Respiratory Society between 2016 and 2019.

| Variables | 2016 | 2017 | 2018 | 2019 | Average yearly change<br>(95%CI), % | P-value | Combined total |
| --- | --- | --- | --- | --- | --- | --- | --- |
| **All pharmaceutical companies** |  |  |  |  |  |  |  |
| **Total payments , \$ (¥)** | 12,439,754<br>(1,355,933,145) | 12,025,229<br>(1,310,749,925) | 13,911,371<br>(1,516,339,453) | 15,171,038<br>(1,653,643,099) | □ | □ | 53,547,391<br>(5,836,665,622) |
| **Average payments±SD, \$** | 4,270±10,590 | 4,368±10,531 | 4,689±10,492 | 5,081±11,520 | 7.6 (5.5□9.8) | <0.001 | 12,134±34,045 |
| **Median payments (IQR), \$** | 1,085<br>(511□3,192) | 1,124<br>(511□3,325) | 1,328<br>(511□3,795) | 1,428<br>(520□4,034) | | | 2,210 (715□8,178) |
| **Payment range, \$** | 85□138,421 | 51□129,078 | 92□149,569 | 31□117,871 | □ | □ | 31□495,332 |
| **Physicians with specific payments, n (%)** |  |  |  |  |  |  |  |
| **Any payments** | 2,913 (40.9) | 2,753 (38.7) | 2,967 (41.7) | 2,986 (42.0) | 1.5 (0.61□2.4) | 0.001 | 4,413 (62.0) |
| **Payments >\$500** | 2,261 (31.8) | 2,174 (30.6) | 2,398 (33.7) | 2,457 (34.5) | 3.6 (2.5□4.6) | <0.001 | 3,747 (52.7) |
| **Payments >\$1,000** | 1,568 (22.0) | 1,490 (20.9) | 1,724 (24.2) | 1,806 (25.4) | 6.0 (4.7□7.3) | <0.001 | 3,000 (42.2) |
| **Payments >\$5,000** | 508 (7.1) | 504 (7.1) | 593 (8.3) | 634 (8.9) | 8.7 (6.3□11.2) | <0.001 | 1478 (20.8) |
| **Payments >\$10,000** | 261 (3.7) | 269 (3.8) | 311 (4.4) | 331 (4.7) | 9.0 (5.7□12.4) | <0.001 | 940 (13.2) |
| **Payments >\$50,000** | 38 (0.53) | 34 (0.48) | 40 (0.56) | 49 (0.69) | 10.2 (-0.76□22.4) | 0.069 | 239 (3.4) |
| **Payments >\$100,000** | 6 (0.084) | 5 (0.070) | 2 (0.028) | 6 (0.084) | -6.1 (-32.5□30.5) | 0.71 | 111 (1.6) |
| **Gini index** | 0.892 | 0.897 | 0.884 | 0.885 | □ | □ | 0.871 |
| **Pharmaceutical companies with 4-years payment data<sup>a</sup>** |  |  |  |  |  |  |  |
| **Total payments , \$** | 12,329,176<br>(1,343,880,199) | 12,010,489<br>(1,309,143,306) | 13,846,437<br>(1,509,261,613) | 15,102,239<br>(1,645,960,030) | □ | □ | 53,286,653<br>(5,808,245,148) |
| **Average payments±SD, \$** | 4,259±10,517 | 4,367±10,519 | 4,679±10,470 | 5,073±11,490 | 7.8 (5.6□9.9) | <0.001 | 12,113±33,943 |
| **Median payments (IQR), \$** | 1,093<br>(511□3,180) | 1,124<br>(511□3,326) | 1,328<br>(511□3,780) | 1,420<br>(520□4,005) | | | 2,214 (715□8,169) |
| **Payment range, \$** | 85□136,230 | 51□129,078 | 92□149,569 | 31□117,871 | □ | □ | 31□495,332 |
| **Physicians with specific payments, n (%)** |  |  |  |  |  |  |  |
| **Any payments** | 2895 (40.7) | 2750 (38.7) | 2959 (41.6) | 2977 (41.8) | 1.6 (0.69□2.5) | 0.001 | 4,399 (61.8) |
| **Payments >\$500** | 2248 (31.6) | 2171 (30.5) | 2391 (33.6) | 2452 (34.5) | 3.7 (2.6□4.7) | <0.001 | 3,734 (52.5) |
| **Payments >\$1,000** | 1562 (22.0) | 1489 (20.9) | 1714 (24.1) | 1803 (25.3) | 6.0 (4.7□7.3) | <0.001 | 2,991 (42.0) |
| **Payments >\$5,000** | 503 (7.1) | 504 (7.1) | 591 (8.3) | 632 (8.9) | 8.9 (6.4□11.4) | <0.001 | 1,472 (20.7) |
| **Payments >\$10,000** | 259 (3.6) | 268 (3.8) | 310 (4.4) | 330 (4.6) | 9.2 (5.9□12.5) | <0.001 | 931 (13.1) |
| **Payments >\$50,000** | 37 (0.52) | 33 (0.46) | 40 (0.56) | 48 (0.67) | 10.7 (-0.36□23.0) | 0.058 | 239 (3.4) |
| **Payments >\$100,000** | 6 (0.084) | 5 (0.070) | 2 (0.028) | 6 (0.084) | -6.1 (-32.5□30.5) | 0.71 | 110 (1.6) |
| **Gini index** | 0.892 | 0.897 | 0.884 | 0.885 | □ | □ | 0.871 |

**IQR: interquartile range; SD: standard deviation**

While 38.0% of the specialists had no personal payments, 52.7%, 42.2%, 20.8%, 13.2%, 3.4%, and 1.6% of respiratory specialists received more than $500, $1,000, $5,000, $10,000, $50,000 and $100,000 in the four-year combined payments, respectively. The Gini index was 0.871 for the four-year combined total payments per specialist, indicating that only a small portion of the respiratory specialists received large amount of the personal payments. Top 1%, 5%, 10% and 25% of specialists occupied 30.3% (95% confidence interval (95% CI): 28.2% □32.4%), 64.8% (95% CI: 62.6% □67.0%), 79.3% (95% CI: 77.8% □80.8%), and 94.2% (95% CI: 93.7% □94.7%) of total monetary values, respectively. (Figure 1) The most common payment category was lecturing, occupying 85.6% of the total ($45,847,076). The average and median payments for top 1% respiratory specialists were $264,879±$70,965 and $247,747 ($209,087□$295,562), respectively. For payment cases per specialist, top 1% specialists received 241.4±67.8 cases in average, and 234 cases in median (195[288) during the four years, respectively.

**Figure 1.**
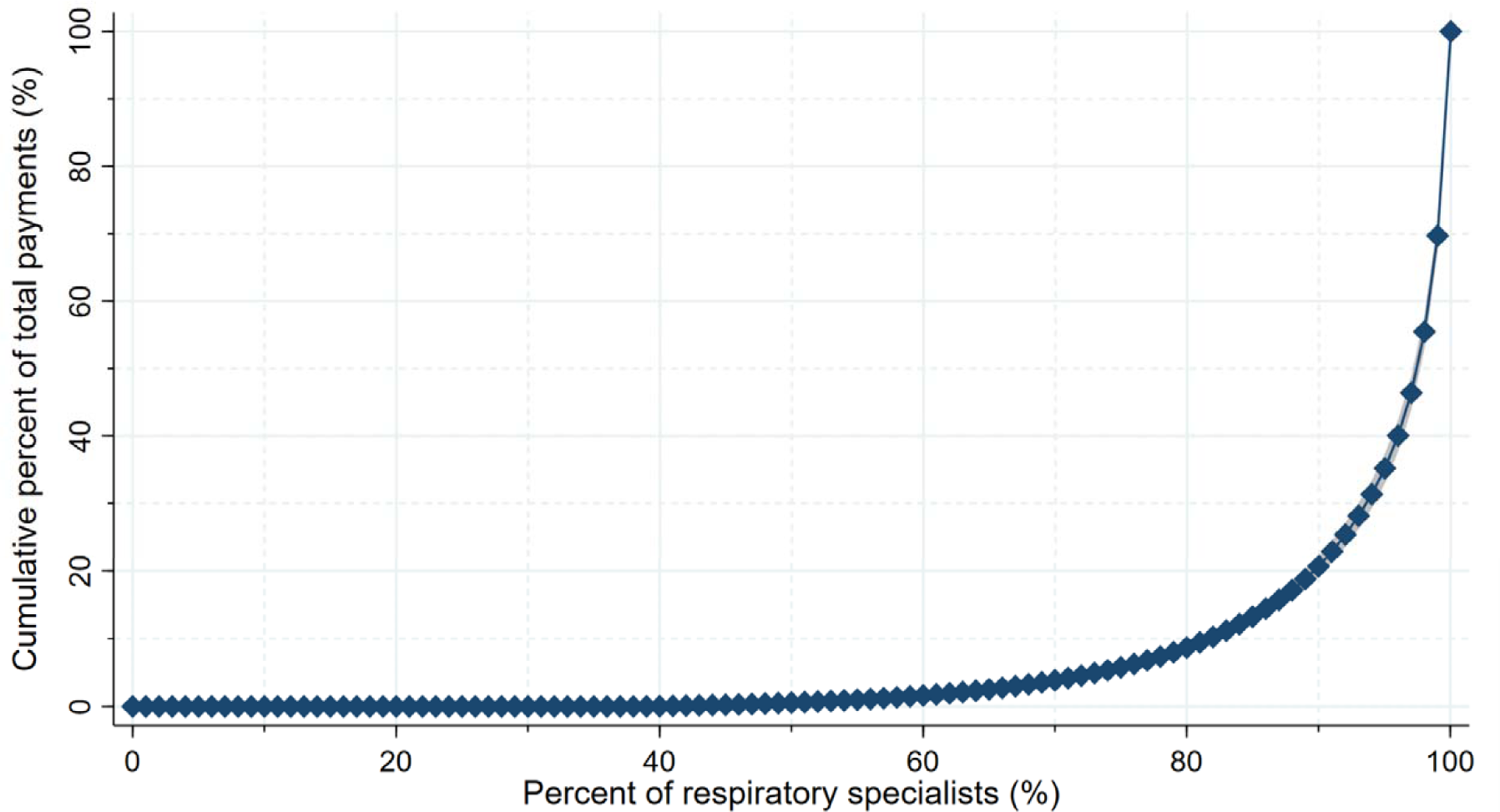
Pharmaceutical payment concentration among the respiratory specialists in Japan.

### Trends of payments

The average payments per specialist ranged $4,270±$10,590 in 2016 to $5,081 ±$11,520 in 2019, respectively. The average annual change rates marked significant increases in both payments per specialist by 7.6% (95% CI: 5.5% □9.8%; p <0.001) and prevalence of specialists with payments by 1.5% (95% CI: 0.61% □2.4%; p = 0.001). Among 72 companies, there were six companies made payments to the specialists without four-year payment data. Therefore, we limited payments from the remaining 66 companies with four-year data. Significant annual increases of payments and prevalence of specialists with payments were also observed, with 7.8% (95% CI: 5.6□9.9; p<0.001) in payments and 1.6% (95%CI: 0.69% □2.5%; p<0.001) in prevalence, respectively.

### Payments by company

Among 72 pharmaceutical companies making payments to the specialists, payments from the top 10 companies accounted for 74.4% of total payments with $39,820,882 between 2016 and 2019. (Figure 2 and Supplemental Material 1) Particularly, the top two companies with the largest amount, AstraZeneca and Boehringer Ingelheim Japan, shared 19.8% ($10,592,516) and 17.6% ($9,428,840) of the total payments. The payment types by top 10 paying companies are shown in Figure 3.

**Figure 2.**
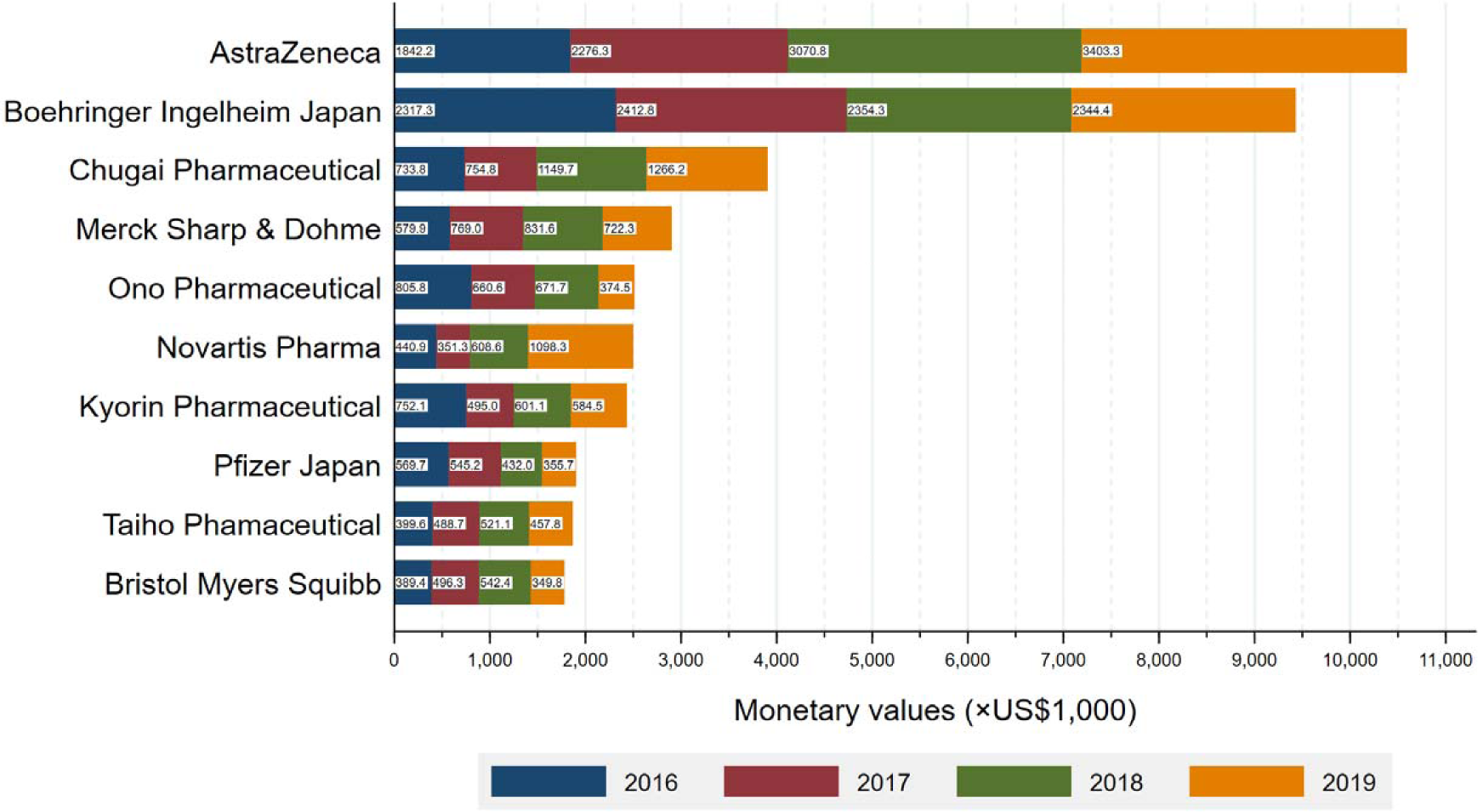
Total payments to respiratory specialists made by the top 10 largest paying companies between 2016 and 2019.

**Figure 3.**
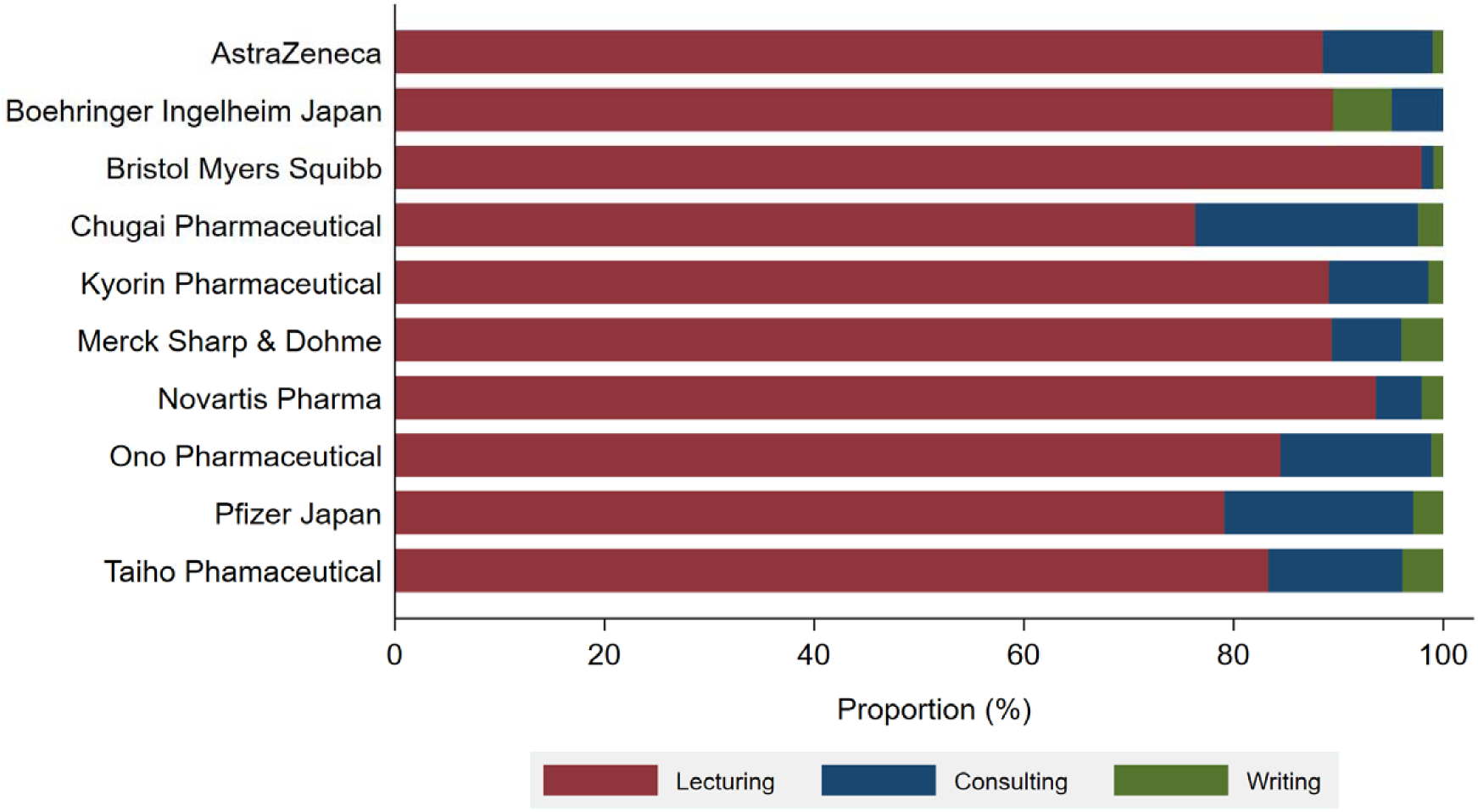
Payment category by the pharmaceutical companies.

### Geographical payment distribution

There were significant geographical differences in the distribution of respiratory and advising specialists in Japan **(Annex 2A and 2B)**. The number of respiratory specialists per million populations ranged from 28.6 in Aomori Prefecture to 86.0 in the Tokyo Metropolis based on the prefecture. The total payment values per prefecture is shown in **Annex** 2C. The average payment values per specialist were the highest in Miyagi Prefecture ($11,855) and the lowest in Oita Prefecture ($3,707) **(Annex 2D)**. For analysis by region, the number of specialists per million populations ranged from 43.8 in Tohoku region (northernmost of Japan) to 64.7 in Kyusyu region (southernmost of Japan), and adversely the average payments per specialist ranged from $6,450 in Kyusyu region to $9,971 in Tohoku region.

## Discussion

Based on the published evidence extracted from Medline/PubMed, Scopus, Web of Science, and Gogle Scholar, this is the first study assessing the magnitude and trends of the financial relationships between respiratory specialists and pharmaceutical companies, which found that 5.4% of total personal payments from all pharmaceutical companies were distributed to respiratory specialists, representing 2.2% (7,114 out of 327,210) of all physicians. Among 7,114 respiratory specialists in Japan, 4,413 (62.0%) accepted $53,547,391 personal payments totaling 74,195 cases from 72 (78.3%) pharmaceutical companies between 2016 and 2019. Further, the respiratory specialists received $12,134 personal payments with 16.8 times for the reimbursement of lecturing, consulting, and writing from 4.8 pharmaceutical companies on average in Japan. These financial interactions significantly increased by 7.8% in payment values per specialist and 1.6% in the prevalence of specialists receiving payments between 2016 and 2019.

First, we should note that there was dearth of research evaluating financial relationships between pharmaceutical companies and respiratory physicians worldwide. However, one study by Inoue et al.^30^ assessed the financial relationships between physicians and pharmaceutical companies across 32 specialties in the US, and reported that pulmonology specialists received an average of $2,689 in general payments such as lecturing, consulting, meals, travel, and accommodations between 2015 and 2017, namely $896 per physician in one year. Comparing with their findings, we observed that the Japanese respiratory specialists received 4.8 to 5.7 times higher average annual payments of $4,270 in 2016, and $5,081 in 2019 ($12,134 in four-year combined average). The discrepancy could have been resulted due to differences in payment categories and a different definition of respiratory specialists between their study in the US and our study in Japan. Nevertheless, our findings indicated strong financial relationships between respiratory physicians and pharmaceutical companies in Japan.

Meanwhile, other studies outside of respiratory specialists reported that there were lower average payments among the US nephrology specialists ($1,795[$2,227 annual general payments)^31^, the US orthopedic surgery ($3,260 in 2013), and the US dermatologists ($4,177)^22^ than the Japanese respiratory specialists; while several specialists such as the US ophthalmologists ($5,191)^32^; the US cardiologists ($5,638)^33^; the US rheumatologists ($6,444)^34^; and Japanese respiratory oncologists ($6,992)^19^ received higher average payments. Particularly, this study represented large financial relationships of JRS-certified specialists with pharmaceutical companies as compared to other specialists in the US and Japan.

One of the primary findings of this study was that both the magnitude and prevalence of payments for respiratory specialists in Japan increased significantly during the past four years, with a 7.8% average annual increase in payments per specialist, and 1.6% in the prevalence of specialists with payments. This finding was consistent with previous studies conducted among US oncologists^28^ and Japanese hematologists. Although Marshall et al. reported that overall payment values and the number of physicians receiving payments declined since the introduction of the US Open Payment Database,^29^ greater payments were increasingly concentrated on a smaller proportion of the US oncologists.^28^ Furthermore, Kusumi et al. reported that both payments and prevalence of hematologists with payments constantly increased with a significant average annual increase of 10.4% and 2.9%, respectively. As Ozaki et al. previously reported, the Japanese respiratory oncologists received the highest personal payments in 2016 due to the launch of the multiple novel oncology drugs for non-small cell lung cancer (NSCLC).^19^

Among the top 10 largest paying companies, in particular, the total annual payments from AstraZeneca, the largest paying company, constantly increased from $1,842,153 in 2016 to $3,403,281 in 2019. Meanwhile, Boehringer Ingelheim, the second largest paying company, remained stable, ranging from $2,317,265 in 2016 to $2,412,839 in 2017. This increasing trend would also be due to the invent of several novel drugs for lung cancers. For example, osimertinib (TAGRISSO® from AstraZeneca) was approved for stage III NSCLC in March 2016, and then it was strongly recommended as a first-line treatment for stage III NSCLC in Japan since 2018.^35^ Consequently, $751 million worth osimertinib was sold in 2019, accounting for the largest share of all oral molecular targeted drugs in Japan.^36^ Also, durvalumab (IMFINZI® from AstraZeneca) received its first approval as an immune checkpoint inhibitor for NSCLC in Japan in July 2018.^35^

Aside from lung cancer drugs, there have been remarkable changes in asthma and chronic obstructive pulmonary disease (COPD) treatment. Although previously monotherapy with inhaled corticosteroids (ICS) was the first-line treatment for COPD, current clinical guidelines such as the Global Initiative for Chronic Obstructive Lung Disease (GOLD) guideline and the JRS guideline recommended dual therapy with the long-acting muscarinic antagonist (LAMA) and long-acting β2-agonist (LABA) or ICS and LABA over monotherapy of ICS or LABA.^37,38^ Following to these changes in guideline recommendations, several companies such as Boehringer Ingelheim, AstraZeneca, and GlaxoSmithKline developed combination drugs of LAMA/LABA, ICS/LABA, and further ICS/LAMA/LABA.^36^ Furthermore, novel biological drugs for asthma treatment have been increasingly developed, such as omalizumab (XOLAIR® from Novartis Pharma, approved for asthma in 2009); mepolizumab (NUCALA® from GlaxoSmithKline, approved asthma and COPD in 2016); benralizumab (FASENRA® from AstraZeneca, approved for asthma in 2018); and duilumab (DUPIXENT® from Sanofi, approved for asthma in 2019).^36^ Considering the recent development of novel drugs in respiratory medicine, the personal payments from pharmaceutical companies to respiratory specialists are expected to increase in Japan in the future.

## Limitations

First, as previously noted, the payment database was structured by the manual collection of payment data from 92 pharmaceutical companies and cross-checked to exclude duplicate physicians from the data. Therefore, this study might have included a few unavoidable human errors, but our repeated and careful scrutiny of payment data by more than two independent reviewers could have minimized such errors. Second, currently, pharmaceutical companies do not disclose their payments concerning meals, beverages, accommodations, travel, and stock ownerships. This might have led to underestimations of the extent and prevalence of overall financial relationships between respiratory specialists and industries. However, such information was not available and the JPMAdoes not mandate to disclose such information on an individual basis.

## Conclusion

The majority of respiratory specialists board certified by the Japanese Respiratory Society increasingly received personal payments for the reimbursements of lecturing, consulting, and writing from the pharmaceutical companies. These financial relationships with pharmaceutical companies were potential COI, and the vast majority of these payments concentrated on only a tiny proportion of specialists. Therefore, we believed this study would reach all respiratory medicine stakeholders, including patients, healthcare professionals, and policymakers, and promote discussion for better healthcare with harmony between patients, healthcare professionals, and industries.

## Data Availability

All data produced in the present study are available upon reasonable request to the authors

## Acknowledgement

The authors thank the Tansa (formerly known as Waseda Chronicle) for providing payment data. Also, we appreciate Mr. Souto Nagano, an undergraduate student from the Faculty of Letters, University of Tokyo; Mr. Kohki Yamada, a medical student at the Osaka University School of Medicine; Mr.Takuto Sakaemura, an undergraduate student from aculty of Applied Science; and Ms. Megumi Aizawa, a graduate student from the Department of Industrial Engineering and Economics, School of Engineering, Tokyo Institute of Technology, for their dedicated contribution on collecting and cross-checking the payment data.

---

**Annex S1.** Value of personal payments by top ten pharmaceutical companies from 2016 to 2019.

**Annex 1. Value of personal payments by top ten pharmaceutical companies from 2016 to 2019**
| Company name | Year, \$ | | | | Combined total, US% (%) |
| --- | --- | --- | --- | --- | --- |
|  | 2016 | 2017 | 2018 | 2019 |  |
| **AstraZeneca** | 1,842,153 | 2,276,273 | 3,070,809 | 3,403,281 | 10,592,516 (19.8) |
| **Boehringer Ingelheim Japan** | 2,317,265 | 2,412,839 | 2,354,322 | 2,344,415 | 9,428,840 (17.6) |
| **Chugai Pharmaceutical** | 733,807 | 754,831 | 1,149,723 | 1,266,167 | 3,904,528 (7.3) |
| **Merck Sharp & Dohme** | 579,941 | 769,027 | 831,594 | 722,307 | 2,902,870 (5.4) |
| **Ono Pharmaceutical** | 805,806 | 660,644 | 671,655 | 374,515 | 2,512,620 (4.7) |
| **Novartis Pharma** | 440,855 | 351,253 | 608,614 | 1,098,319 | 2,499,041 (4.7) |
| **Kyorin Pharmaceutical** | 752,102 | 495,036 | 601,077 | 584,490 | 2,432,705 (4.5) |
| **Pfizer Japan** | 569,670 | 545,174 | 432,017 | 355,677 | 1,902,539 (3.6) |
| **Taiho Pharmaceutical** | 399,571 | 488,742 | 521,074 | 457,771 | 1,867,159 (3.5) |
| **Bristol Myers Squibb** | 389,440 | 496,347 | 542,435 | 349,841 | 1,778,064 (3.3) |

**Annex 2A:**
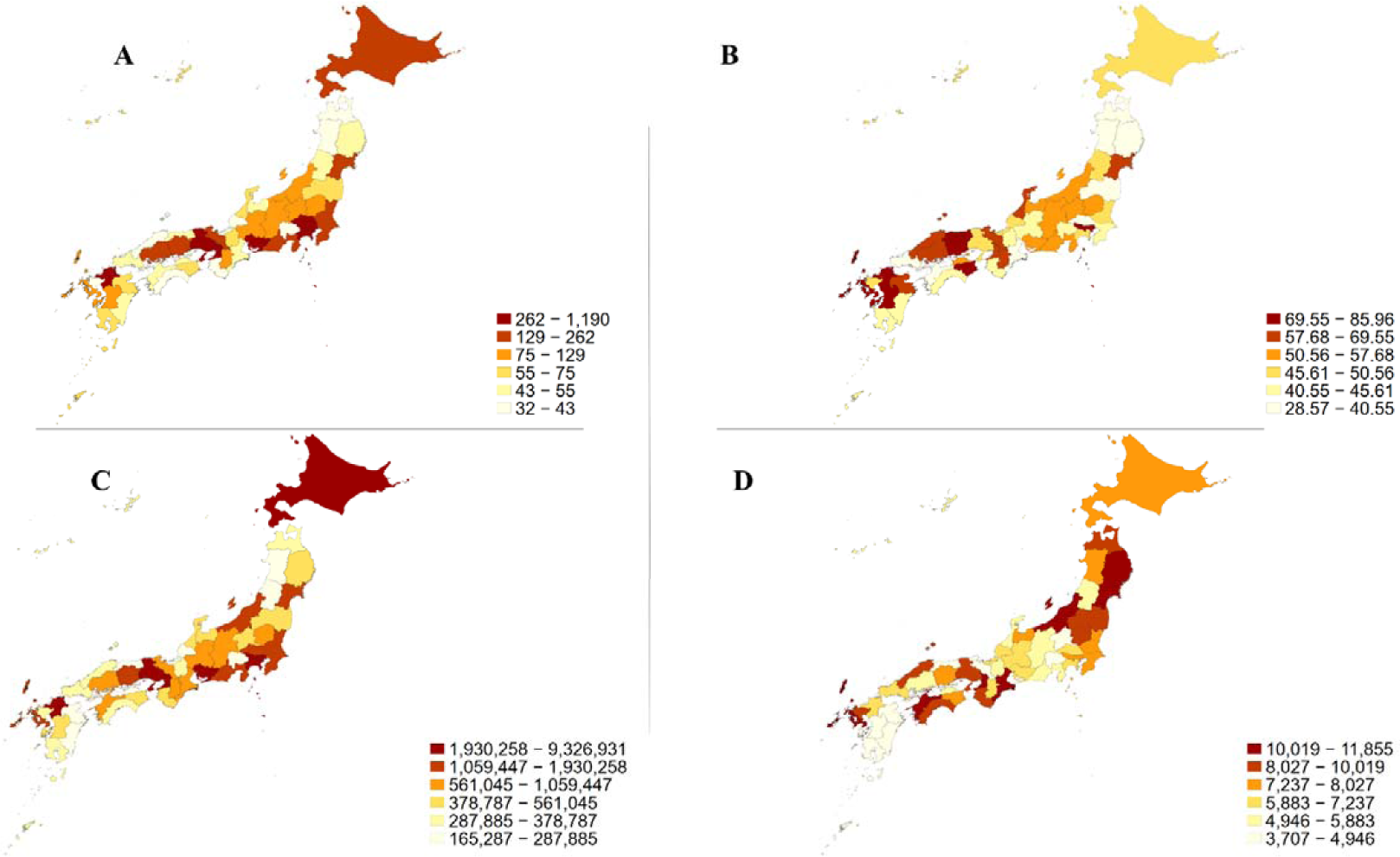
Geographical differences in number of respiratory specialists per million population in 2021; 2B: number of teaching respiratory specialists per million population in 2021; 2C: total personal payment values from 2016 to 2019; 2D: average personal payment values per respiratory specialist from 2016 to 2019

